# Corowa-kun: Impact of a COVID-19 vaccine information chatbot on vaccine hesitancy, Japan 2021

**DOI:** 10.1101/2021.05.26.21257854

**Authors:** Takaaki Kobayashi, Yuka Nishina, Hana Tomoi, Ko Harada, Kyuto Tanaka, Eiyu Matsumoto, Kenta Horimukai, Jun Ishihara, Shugo Sasaki, Kanako Inaba, Kyosuke Seguchi, Hiromizu Takahashi, Jorge L. Salinas, Yuji Yamada

## Abstract

**Background:** Few studies have assessed how mobile messenger apps affect COVID-19 vaccine hesitancy. We created a COVID-19 vaccine information chatbot in a popular messenger app in Japan to answer commonly asked questions.

**Methods:** LINE is the most popular messenger app in Japan. Corowa-kun, a free chatbot, was created in LINE on February 6, 2021. Corowa-kun provides instant, automated answers to frequently asked COVID-19 vaccine questions. In addition, a cross-sectional survey assessing COVID-19 vaccine hesitancy was conducted via Corowa-kun during April 5–12, 2021.

**Results:** A total of 59,676 persons used Corowa-kun during February–April 2021. Of them, 10,192 users (17%) participated in the survey. Median age was 55 years (range 16–97), and most were female (74%). Intention to receive a COVID-19 vaccine increased from 59% to 80% after using Corowa-kun (p < 0.01). Overall, 20% remained hesitant: 16% (1,675) were unsure, and 4% (364) did not intend to be vaccinated. Factors associated with vaccine hesitancy were: age 16 to 34 (odds ratio [OR] = 3.7, 95% confidential interval [CI]: 3.0–4.6, compared to age ≥65), female sex (OR = 2.4, Cl: 2.1–2.8), and history of a previous vaccine side-effect (OR = 2.5, Cl: 2.2–2.9). Being a physician (OR = 0.2, Cl: 0.1–0.4) and having received a flu vaccine the prior season (OR = 0.4, Cl: 0.3–0.4) were protective.

**Conclusions:** Corowa-kun reduced vaccine hesitancy by providing COVID-19 vaccine information in a messenger app. Mobile messenger apps could be leveraged to increase COVID-19 vaccine acceptance.

## Text

### Background

In 2020, a global coronavirus disease-2019 (COVID-19) pandemic emerged, caused by the SARS-CoV-2 virus. [1] According to the World Health Organization, there have been 134 million confirmed cases of COVID-19, including 2.9 million deaths, as of April 8, 2021. [2] Multiple COVID-19 vaccines are highly effective at preventing symptomatic disease. [3] Most developed countries have already vaccinated large proportions of their population. However, many individuals choose not to be vaccinated, often citing safety concerns. Studies in multiple settings have revealed high levels of COVID-19 vaccine hesitancy, ranging from 20–40% of the surveyed populations. [4-8] Vaccine hesitancy differs depending on sociodemographic factors, such as race and educational level, as well as attitudes and beliefs. [9-13] Understanding peoples’ concerns about COVID-19 vaccines is necessary to increase vaccine uptake among those who are hesitant.

Japan has one of the highest vaccine hesitancy rates in the world. According to a previous study, less than 30% of people strongly agreed that vaccines were safe, important, or effective. [14] Moreover, Japan has a long history of public concerns about vaccine adverse events. The measles, mumps, and rubella (MMR) vaccine was introduced into the national immunization program in 1989. However, because of reports of aseptic meningitis following the MMR vaccine, the Japanese government withdrew its recommendation for the MMR vaccine in 1993. [15] Human papillomavirus (HPV) vaccination for girls ages 12–16 was first licensed in Japan in October 2009. Acceptance initially reached over 70%, but it fell to less than 1% after extensive media reports of possible adverse events. [16] Ultimately, the Japanese government suspended its recommendation for the vaccine in June 2013. [17]

Social media are popular in Japan, allowing for circulation of user-generated content and social interaction. Using social media is an integral part of daily life for many people. Leveraging social media platforms to promote vaccines has been investigated. One study revealed that mothers presented with vaccine information on social media during their pregnancy were more likely to vaccinate their infants on time. [18] However, there is a paucity of studies assessing the effect of messenger apps on COVID-19 vaccine acceptance.

We created a chatbot in an already existing popular messenger app in Japan to provide people with COVID-19 vaccine information via text messages. We assessed the impact of chatbot text messages on COVID-19 vaccine hesitancy by conducting a cross-sectional survey among chatbot users.

## Methods

### Corowa-kun: A chatbot with COVID-19 vaccine information in LINE

LINE is a free messenger app available for use in electronic devices, such as smartphones, tablets, and personal computers. LINE users can exchange texts, images, video, and audio. LINE is the most popular messenger app in Japan; about 86 million people in Japan (roughly two-thirds of the population) use this messenger app. [19]

We created a chatbot in LINE called Corowa-kun’s Consultation Room (Corowa-kun) (Figure 1 and supplemental figure 1) to answer COVID-19 vaccine frequently asked questions (FAQs) via text messages. Corowa-kun is free, and anyone using LINE can access it. To create Corowa-kun, we first searched for COVID-19 vaccine FAQs using Japanese government websites and the United States Centers for Disease Control and Prevention website. [20] [21] Fifty questions that we thought were important were selected. We then composed our own corresponding answers to these fifty questions and used them as the initial content for Corowa-kun. Corowa-kun went live in LINE on February 6, 2021. Users have the option to enter free-text questions, and twenty additional FAQs with answers were added based on users’ free-text questions entered by April 5, 2021. The public was made aware of Corowa-kun using mass media (e.g. television, radio, newspapers). [22]

**Figure 1:**
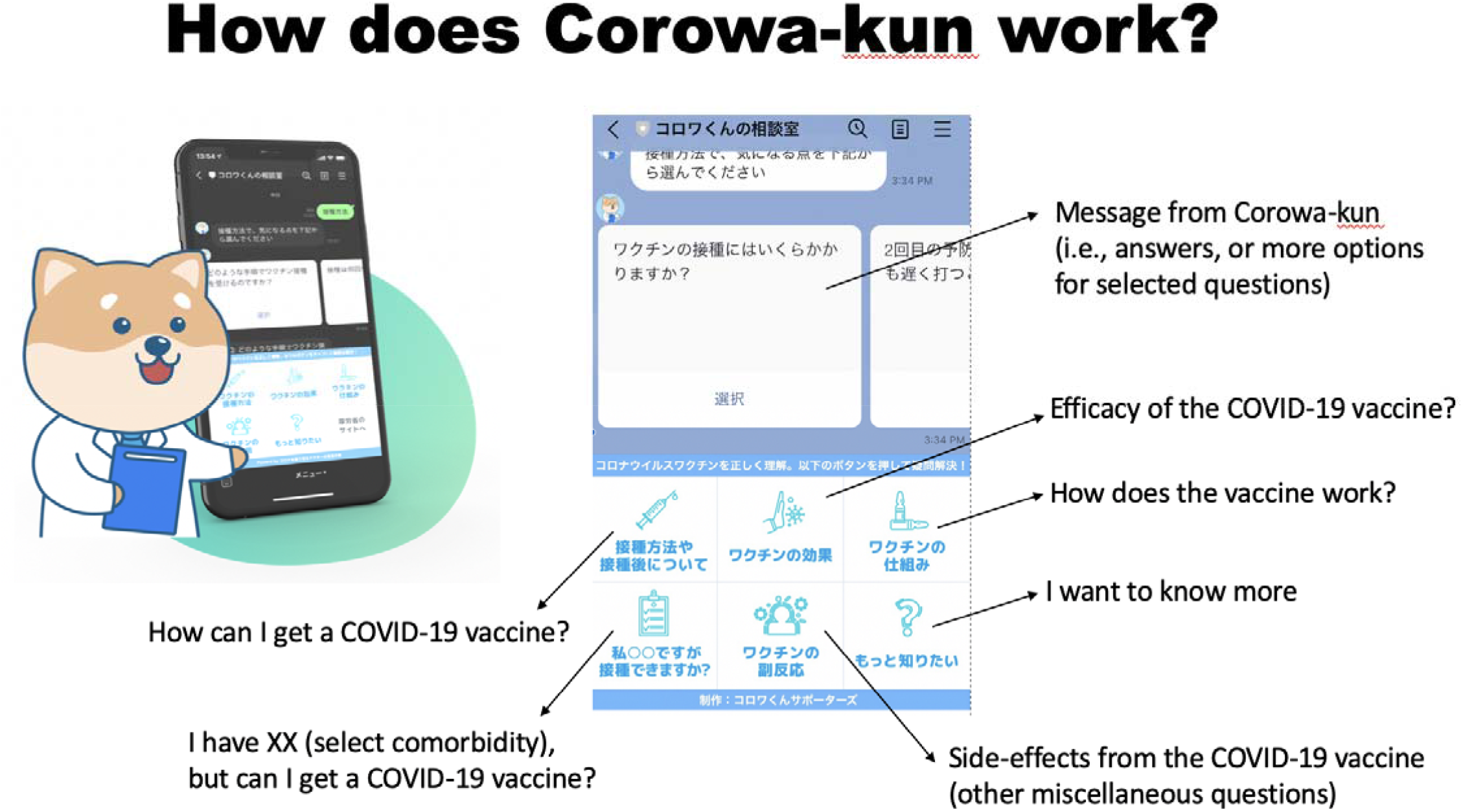
Corowa-kun’s Consultation Room: a free messenger app chatbot, Japan 2021. Corowa-kun is the mascot of an online chatbot. This chatbot in LINE is used to answer COVID-19 vaccine frequently asked questions (FAQs) via text messages. As of May 10^th^, 70 FAQs are available.

### Corowa-kun users and frequently accessed FAQs

We tracked the total number of users who accessed and used Corowa-kun from February 6 to April 12, 2021. We analyzed how frequently each FAQ was accessed from April 5 to 12, 2021. We did not examine frequency of FAQ access before April 5 because new FAQ messages were added prior to then, and the total number of FAQs changed from the initial content in February.

### Cross-sectional survey

A cross-sectional survey was conducted using Corowa-kun during April 5–12, 2021. All persons who accessed Corowa-kun by April 12, 2021 were invited within Corowa-kun to participate in the survey. We did not use any incentives, and the survey was completely voluntary. The survey started with three screening questions: (1) “Are you 16 years old or older?”; (2) “Have you received a COVID-19 vaccine?”; and, (3) “Do you agree to participate?”. We included persons ages 16 years old and older who had not received a COVID-19 vaccine. Those who agreed to participate received a link to a Google form. Each LINE account could only answer the survey once.

The survey was written in Japanese and consisted of 21 questions. To examine attitudes and beliefs regarding COVID-19 vaccines, we included survey items used in similar studies and added our own questions (supplemental document 1). [4] We asked for age, sex, geographic location, educational attainment, employment status, occupation, marriage status, pregnancy status, household members <16 or >64 years old, household size, annual household income, presence of chronic diseases identified as risk factors for severe COVID-19 [21], smoking status, history of influenza vaccine in the previous season, self-rated overall health (scale from 1 to 9 with 9 being best), history of COVID-19, whether they would like to get COVID-19 vaccination once available (“I would like to get the vaccine” = Yes, “I am not sure” = Unsure, and “I do not want to get the vaccine” = No). Geographic locations were combined using the following categories: Hokkaido, Tohoku, Kanto (e.g., Tokyo), Chubu, Kansai (e.g., Osaka), Kinki, Chugoku, Shikoku, Kyushu regions, and outside Japan. [23] We asked why they did or did not want to get COVID-19 vaccines using pre-defined answers with a free-text comment (supplemental document 1, respondents could choose multiple answers). Since we aimed to investigate the impact of Corowa-kun on COVID-19 vaccine hesitancy, we also asked whether there was any change in intent to be vaccinated before and after using Corowa-kun.

### Statistical analysis

Participant characteristics were summarized using frequencies and percentages. For two group comparisons, the Chi-squared test was used for categorical variables and the U-Mann Whitney test was used for continuous variables. For three group comparisons, the Chi-squared test was used for categorical variables. To identify risk factors for vaccine hesitancy, those responding “No” and “Unsure” were combined (vaccine hesitant) and compared to those responding “Yes”. We fit separate univariate logistic regression models for each exposure, limited to groups with 4 or more persons. We used SAS (version 9, Carey, NC) for statistical analysis. A p-value of 0.05 was considered statistically significant. This study was approved by the institutional review board of Kanto Central Hospital.

## Results

A total of 59,676 persons used Corowa-kun by April 12, 2021. During the cross-sectional survey period (April 5–12, 2021), a total of 14,240 FAQ text messages were used. The most commonly accessed message categories were: “I have (select comorbidity), can I get a COVID-19 vaccine?” (23%); followed by questions on adverse reactions (22%), and how the vaccine works (20%).

Of 59,676 users, 2,472 had the LINE option to block surveys turned on or had inactive account, and 57,235 received the survey invitation. Of those who received it, 10,331 (18.1%) responded to the screening questions. Of those who responded, 139 users were excluded: 26 users were age <16, and 113 users had already received a COVID-19 vaccine. A total of 10,192 (17.1%) were included in the survey analysis. Median age was 55 years (range 16–97), and most were women (74%, table 1). The most common respondent regions were Kansai (42.4%) and Kanto (31.4%, supplemental figure 2). Almost all participants (98%) had a high school diploma or more advanced education. Healthcare workers consisted of 18% of participants: 171 (1.7%) were physicians and 1,723 (16.9%) were healthcare workers other than physicians. Sixty-six participants (0.7%) were pregnant. Participants who lived with someone age ≤16 comprised 19.2% of the study population, and those who lived with someone age ≥65 were 35.7%. The most common household size was three (25.5%).

**Table 1:** Characteristics of the COVID-19 vaccine survey participants, Japan, 2021 (n=10,192)

|  | Total | Intend to get vaccinated | Do not intend to get vaccinated | Unsure |
| --- | --- | --- | --- | --- |
| Characteristics | N=10,192 | N= 8,153 | N= 364 | N= 1,675 |
| **Sociodemographic** |  |  |  |  |
| **Age** |  |  |  |  |
| Mean (SD) | 54 (12) | 54.6 (12.4) | 48.6 (12.8) | 50.6 (11.4) |
| **Age category** |  |  |  |  |
| 16-34 | 798 (7.8%) | 569 (7.0%) | 61 (16.8%) | 168 (10.0%) |
| 35-49 | 2694 (26.4%) | 2046 (25.1%) | 122 (33.5%) | 526 (31.4%) |
| 50-64 | 4562 (44.8%) | 3609 (44.3%) | 143 (39.3%) | 810 (48.4%) |
| ≥65 | 2138 (21.0%) | 1929 (23.7%) | 38 (10.4%) | 171 (10.2%) |
| **Sex** |  |  |  |  |
| Male | 2556 (25.1%) | 2778 (27.9%) | 62 (17.0%) | 216 (12.9%) |
| Female | 7543 (74.0%) | 5816 (71.3%) | 292 (80.2%) | 1435 (85.7%) |
| Other | 17 (0.2%) | 14 (0.2%) | 1 (0.3%) | 2 (0.1%) |
| No answer | 76 (0.8%) | 45 (0.6%) | 9 (2.5%) | 22 (1.3%) |
| **Region** |  |  |  |  |
| Hokkaido | 270 (2.7%) | 198 (2.4%) | 15 (4.1%) | 57 (3.4%) |
| Tohoku | 315 (3.1%) | 236 (2.9%) | 11 (3.0%) | 68 (4.1%) |
| Kanto | 3203 (31.4%) | 2606 (32.0%) | 118 (32.4%) | 479 (28.6%) |
| Chubu | 1091 (10.7%) | 868 (10.7%) | 38 (10.4%) | 185 (11.0%) |
| Kansai | 4325 (42.4%) | 3497 (42.9%) | 125 (34.3%) | 703 (42.0%) |
| Chugoku | 271 (2.7%) | 214 (2.6%) | 14 (3.9%) | 43 (2.6%) |
| Shikoku | 156 (1.5%) | 109 (1.3%) | 9 (2.5%) | 38 (2.3%) |
| Kyusyu | 527 (5.2%) | 399 (4.9%) | 31 (8.5%) | 97 (5.8%) |
| Abroad | 34 (0.3%) | 26 (0.3%) | 3 (0.8%) | 5 (0.3%) |
| **Educational attainment** |  |  |  |  |
| Elementary or junior high school | 208 (2.0%) | 151 (1.9%) | 10 (2.8%) | 47 (2.8%) |
| High school | 2688 (26.2%) | 2008 (24.6%) | 119 (32.7%) | 541 (32.3%) |
| College or professional school | 3365 (33.0%) | 2635 (32.3%) | 121 (33.2%) | 609 (36.4%) |
| Undergraduate school | 3502 (34.4%) | 2966 (36.4%) | 100 (27.5%) | 436 (26.0%) |
| Postgraduate school | 449 (4.4%) | 393 (4.8%) | 14 (3.9%) | 42 (2.5%) |
| **Employment status** |  |  |  |  |
| Full-time | 3866 (37.9%) | 3142 (38.5%) | 142 (39.0%) | 582 (34.8%) |
| Part-time | 2728 (26.8%) | 2097 (25.7%) | 114 (31.3%) | 517 (30.9%) |
| Student | 100 (1.0%) | 73 (0.9%) | 3 (0.8%) | 24 (1.4%) |
| Retired | 740 (7.3%) | 681 (8.4%) | 9 (2.5%) | 50 (3.0%) |
| Homemaker | 1992 (19.5%) | 1555 (19.1%) | 69 (19.0%) | 368 (22.0%) |
| Unemployed due to COVID-19 | 128 (1.3%) | 91 (1.1%) | 6 (1.7%) | 31 (1.9%) |
| Unemployed irrelevant to COVID-19 | 638 (6.3%) | 514 (6.3%) | 21 (5.8%) | 103 (6.2%) |
| **Healthcare worker** |  |  |  |  |
| Physician | 171 (1.7%) | 164 (2.0%) | 3 (0.8%) | 4 (0.2%) |
| Yes, other than physician | 1723 (16.9%) | 1400 (17.2%) | 75 (20.6%) | 248 (14.8%) |
| No | 8298 (81.4%) | 6589 (80.8%) | 286 (78.6%) | 1423 (85.0%) |
| **Marriage status** |  |  |  |  |
| Married | 7412 (72.7%) | 6038 (74.1%) | 218 (59.9%) | 1156 (69.0%) |
| Never married | 1841 (18.1%) | 1409 (17.3%) | 94 (25.8%) | 338 (20.2%) |
| Divorced | 939 (9.2%) | 706 (8.7%) | 52 (14.3%) | 181 (10.8%) |
| **Living with persons at age<16** | 1958 (19.2%) | 1506 (18.5%) | 81 (22.3%) | 371 (22.2%) |
| **Living with persons at age≥65** | 3641 (35.7%) | 2994 (36.7%) | 112 (30.8%) | 535 (31.9%) |
| **Household size** |  |  |  |  |
| 1 | 1499 (14.8%) | 1199 (14.8%) | 65 (18.2%) | 235 (14.2%) |
| 2 | 3685 (36.5%) | 3025 (37.4%) | 119 (33.3%) | 541 (32.6%) |
| 3 | 2577 (25.5%) | 2009 (24.8%) | 88 (24.7%) | 480 (29.0%) |
| 4 | 1576 (15.6%) | 1266 (15.7%) | 52 (14.6%) | 258 (15.6%) |
| 5 | 500 (5.0%) | 383 (4.7%) | 21 (5.9%) | 96 (5.8%) |
| ≥6 | 265 (2.6%) | 205 (2.5%) | 12 (3.4%) | 48 (2.9%) |
| **Annual household income** |  |  |  |  |
| Less than JPY 2 million | 4592 (45.1%) | 3503 (43.0%) | 160 (44.0%) | 929 (55.5%) |
| JPY 200 million < JPY 400 million | 2823 (27.7%) | 2286 (28.0%) | 110 (30.2%) | 427 (25.5%) |
| JPY 400 million < JPY 600 million | 1449 (14.2%) | 1214 (14.9%) | 56 (15.4%) | 179 (10.7%) |
| JPY 600 million < JPY 800 million | 664 (6.5%) | 560 (6.9%) | 21 (5.8%) | 83 (5.0%) |
| JPY 800 million or higher | 664 (6.5%) | 590 (7.2%) | 17 (4.7%) | 57 (3.4%) |
| **Health status** |  |  |  |  |
| **Chronic respiratory disease** | 795 (7.8%) | 588 (7.2%) | 41 (11.3%) | 166 (9.9%) |
| **Chronic heart disease** | 1419 (13.9%) | 1190 (14.6%) | 31 (8.5%) | 198 (11.8%) |
| **Chronic kidney disease** | 152 (1.5%) | 130 (1.6%) | 6 (1.7%) | 16 (1.0%) |
| **Chronic liver disease** | 84 (0.8%) | 70 (0.9%) | 2 (0.6%) | 12 (0.7%) |
| **Diabetes mellitus** | 529 (5.2%) | 439 (5.4%) | 15 (4.1%) | 75 (4.5%) |
| **Blood disease except for anaemia** | 154 (1.5%) | 119 (1.5%) | 9 (2.5%) | 26 (1.6%) |
| **Malignancy** | 216 (2.1%) | 149 (1.8%) | 17 (4.7%) | 50 (3.0%) |
| **Receiving immunosuppressant** | 309 (3.0%) | 224 (2.8%) | 14 (3.9%) | 71 (4.2%) |
| **Neurological or neuromuscular disease due to immune deficiency** | 60 (0.6%) | 36 (0.4%) | 5 (1.4%) | 19 (1.1%) |
| **Physical decline associated with a neurological disease or a neuromuscular disease** | 34 (0.3%) | 23 (0.3%) | 1 (0.3%) | 10 (0.6%) |
| **Chromosomal abnormality** | 14 (0.1%) | 11 (0.1%) | 1 (0.3%) | 2 (0.1%) |
| **Severe psychosomatic disorder** | 7 (0.07%) | 3 (0.04%) | 2 (0.6%) | 2 (0.1%) |
| **Sleep apnoea** | 271 (2.7%) | 224 (2.8%) | 11 (3.0%) | 36 (2.2%) |
| **Obesity** | 936 (9.2%) | 704 (8.6%) | 24 (6.6%) | 208 (12.4%) |
| **No underlying health condition** | 5862 (57.5%) | 4767 (58.5%) | 219 (60.2%) | 876 (52.3%) |
| **Smoking status** |  |  |  |  |
| Active smoker | 922 (9.1%) | 690 (8.5%) | 47 (12.9%) | 185 (11.0%) |
| Former smoker | 2435 (23.9%) | 1987 (24.4%) | 83 (22.8%) | 365 (21.8%) |
| Only electronic cigarettes | 278 (2.7%) | 237 (2.9%) | 8 (2.2%) | 33 (2.0%) |
| Never smoker | 6557 (64.3%) | 5239 (64.3%) | 226 (62.1%) | 1092 (65.2%) |
| **Have you had a flu shot within the past year?** | 7037 (69.0%) | 5998 (73.6%) | 131 (36.0%) | 908 (54.2%) |
| **Self-reported history of COVID-19** |  |  |  |  |
| Yes (I tested positive) | 58 (0.6%) | 48 (0.6%) | 3 (0.8%) | 7 (0.4%) |
| Yes (I had the symptoms but did not receive a positive test) | 83 (0.8%) | 60 (0.7%) | 5 (1.4%) | 18 (1.1%) |
| No | 10051 (98.6%) | 8045 (98.7%) | 356 (97.8%) | 1650 (98.5%) |
| **Have you had any vaccine side-effects?** |  |  |  |  |
| Yes | 975 (9.6%) | 644 (7.9%) | 85 (23.4%) | 246 (14.7%) |
| No | 8475 (83.2%) | 7051 (86.5%) | 238 (65.4%) | 1186 (70.8%) |
| Unsure | 742 (7.3%) | 458 (5.6%) | 41 (11.3%) | 243 (14.5%) |
| **Self-reported overall health** |  |  |  |  |
| 1-3 | 116 (1.1%) | 76 (0.9%) | 11 (3.0%) | 29 (1.7%) |
| 4-6 | 5430 (53.3%) | 4144 (50.8%) | 222 (61.0%) | 1064 (63.5%) |
| 7-9 | 4646 (45.6%) | 3933 (48.2%) | 131 (36.0%) | 582 (34.8%) |
| **Pregnancy status** |  |  |  |  |
| Pregnant | 67 (0.7%) | 36 (0.4%) | 5 (1.4%) | 26 (1.6%) |
| Not pregnant | 5159 (50.6%) | 4087 (50.1%) | 176 (48.4%) | 896 (53.5%) |
| Desire to be pregnant | 228 (2.2%) | 144 (1.8%) | 26 (7.1%) | 58 (3.5%) |
| Not applicable | 4738 (46.5%) | 3886 (47.7%) | 157 (43.1%) | 695 (41.5%) |

Figure 2 displays intent for being vaccinated before and after use of Corowa-kun. Participants who intended to be vaccinated increased after using Corowa-kun (p < 0.01). Overall, after use of Corowa-kun, 80.0% of participants intended to be vaccinated (an increase from 59% before use), 16.4% were not sure, and 3.6% did not intend to be vaccinated.

**Figure 2:**
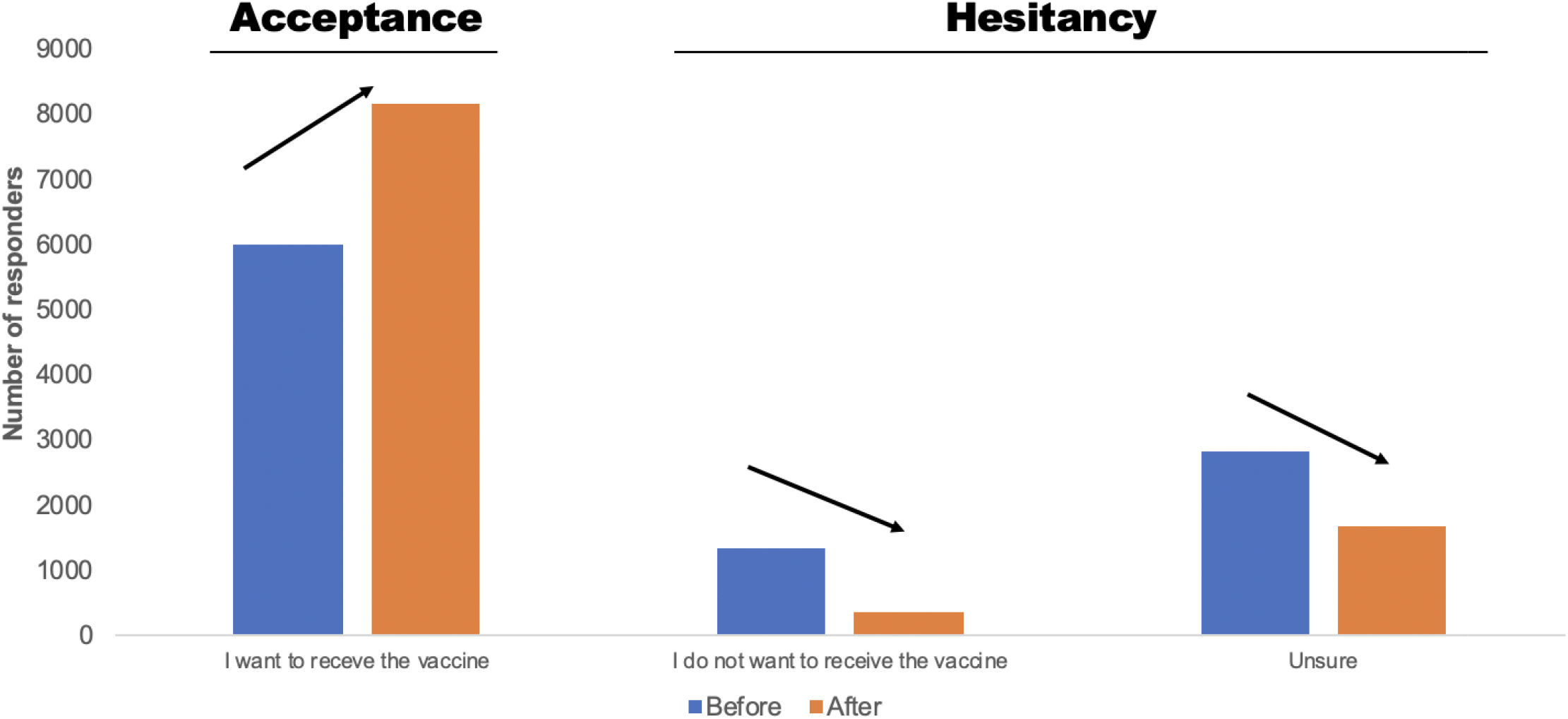
COVID-19 vaccine acceptance increased and hesitancy decreased after using Corowa-kun, Japan, 2021 (n=10,192) *There was a statistically significant difference in responses between before and after using Corowa-kun (p < 0.01, Chi-square test)

Participants who did not intend to be vaccinated or were not sure were combined (vaccine hesitant; n= 2,039) and compared to participants who intended to be vaccinated (n=8,153). Table 2 summarizes the factors associated with vaccine hesitancy: Age 16 to 34 (odds ratio [OR] = 3.7, 95% confidential interval [CI]: 3.0–4.6, compared to age ≥65), female sex (OR = 2.4, Cl: 2.1–2.8), pregnancy (OR = 3.3, Cl: 2.0–5.3), and history of vaccine side-effects (OR = 2.5, Cl: 2.2–2.9). Being a physician (OR = 0.2, Cl: 0.1–0.4, compared to non-healthcare workers) and receiving a flu vaccine in the last influenza season (OR = 0.4, Cl: 0.3–0.4) were protective.

**Table 2:** Univariable logistic regression models of factors associated with COVID-19 vaccine hesitancy, Japan, 2021 (n=10,192)

|  | Vaccine hesitancy<br>N=2,039 |  | Vaccine acceptance<br>N=8,153 |  | Odds ratio |
| --- | --- | --- | --- | --- | --- |
| Age |  |  |  |  |  |
| 16-34 | 229 | 11.2% | 569 | 7.0% | 3.7 (3.0-4.6) |
| 35-49 | 648 | 31.8% | 2046 | 25.1% | 2.9 (2.5-3.5) |
| 50-64 | 953 | 46.7% | 3609 | 44.3% | 2.4 (2.1-2.9) |
| ≥65 | 209 | 10.3% | 1929 | 23.7% | Ref |
| Sex |  |  |  |  |  |
| Male | 278 | 13.63% | 2278 | 27.9% | Ref |
| Female | 1727 | 84.7% | 5816 | 71.3% | 2.4 (2.1-2.8) |
| Other | 3 | 0.2% | 14 | 0.2% | NA |
| No answer | 31 | 1.5% | 45 | 0.6% | 5.6 (3.5-9.1) |
| Region |  |  |  |  |  |
| Hokkaido | 72 | 3.5% | 198 | 2.4% | 1.6 (1.2-2.1) |
| Tohoku | 79 | 3.9% | 236 | 2.9% | 1.5 (1.1-1.9) |
| Kanto | 597 | 29.3% | 2606 | 32.0% | Ref |
| Chubu | 223 | 10.9% | 868 | 10.7% | 1.1 (0.9-1.3) |
| Kansai | 828 | 40.6% | 3497 | 42.9% | 1.0 (0.9-1.2) |
| Chugoku | 57 | 2.8% | 214 | 2.6% | 1.2 (0.9-1.6) |
| Shikoku | 47 | 2.3% | 109 | 1.3% | 1.8 (1.3-2.6) |
| Kyusyu | 128 | 6.3% | 399 | 1.9% | 1.4 (1.1-1.7) |
| Abroad | 8 | 0.4% | 26 | 0.3% | 1.3 (0.6-3.0) |
| Educational attainment |  |  |  |  |  |
| Elementary or junior high school | 57 | 2.8% | 151 | 1.9% | 1.4 (1.0-1.9) |
| High school | 660 | 32.4% | 2008 | 24.6% | 1.2 (1.1-1.3) |
| College or professional school | 730 | 35.8% | 2635 | 32.3% | Ref |
| Undergraduate school | 536 | 26.3% | 2966 | 36.4% | 0.7 (0.6-0.7) |
| Postgraduate school | 56 | 2.8% | 393 | 4.8% | 0.5 (0.4-0.7) |
| Employment status |  |  |  |  |  |
| Full-time | 724 | 35.5% | 3142 | 38.5% | Ref |
| Part-time | 631 | 31.0% | 2097 | 25.7% | 1.3 (1.2-1.5) |
| Student | 27 | 1.3% | 73 | 0.9% | 1.6 (1.0-2.5) |
| Retied | 59 | 2.9% | 681 | 8.4% | 0.4 (0.3-0.5) |
| Homemaker | 437 | 21.4% | 1555 | 19.1% | 1.2 (1.1-1.4) |
| Unemployed due to COVID-19 | 37 | 1.8% | 91 | 1.1% | 1.8 (1.2-2.6) |
| Unemployed irrelevant to COVID-19 | 124 | 6.1% | 514 | 6.3% | 1.0 (0.8-1.3) |
| Healthcare worker |  |  |  |  |  |
| Physician | 7 | 0.3% | 164 | 2.0% | 0.2 (0.1-0.4) |
| Yes, but not physician | 323 | 15.8% | 1400 | 17.2% | 0.9 (0.8-1.01) |
| No | 1709 | 83.8% | 6589 | 80.8% | Ref |
| Marriage status |  |  |  |  |  |
| Married | 1374 | 67.4% | 6038 | 47.1% | Ref |
| Never married | 432 | 21.2% | 1409 | 17.3% | 1.3 (1.2-1.5) |
| Divorced | 233 | 11.4% | 706 | 8.7% | 1.5 (1.2-1.7) |
| Living with persons at age<16 | 452 | 22.2% | 1506 | 18.5% | 1.3 (1.1-1.4) |
| Living with persons at age≥65 | 647 | 31.8% | 2994 | 36.7% | 0.8 (0.7-0.9) |
| Household size |  |  |  |  |  |
| 1 | 300 | 14.9% | 1199 | 14.8% | Ref |
| 2 | 660 | 32.8% | 3025 | 37.4% | 0.9 (0.7-1.0) |
| 3 | 568 | 28.2% | 2009 | 24.8% | 1.1 (1.0-1.3) |
| 4 | 310 | 15.4% | 1266 | 15.7% | 1.0 (0.8-1.2) |
| 5 | 117 | 5.8% | 383 | 4.8% | 1.2 (1.0-1.6) |
| ≥6 | 60 | 3.0% | 205 | 2.5% | 1.2 (0.9-1.6) |
| Annual household income |  |  |  |  |  |
| Less than JPY 2 million | 1089 | 53.4% | 3503 | 43.0% | 1.6 (1.4-1.9) |
| JPY 200 million < JPY 400 million | 537 | 26.3% | 2286 | 28.0% | 1.2 (1.0-1.4) |
| JPY 400 million < JPY 600 million | 235 | 11.5% | 1214 | 14.9% | Ref |
| JPY 600 million < JPY 800 million | 104 | 5.1% | 560 | 6.9% | 1.0 (0.7-1.2) |
| JPY 800 million or higher | 74 | 3.6% | 590 | 7.2% | 0.7 (0.5-0.9) |
| Chronic respiratory disease | 207 | 10.2% | 588 | 7.2% | 1.5 (1.2-1.7) |
| Chronic heart disease | 229 | 11.2% | 1190 | 14.6% | 0.7 (0.6-0.9) |
| Chronic kidney disease | 22 | 1.1% | 130 | 1.6% | 0.7 (0.4-1.1) |
| **Chronic liver disease** | 14 | 0.7% | 70 | 0.9% | 0.8 (0.4-1.4) |
| **Diabetes mellitus** | 90 | 4.4% | 439 | 5.4% | 0.8 (0.6-1.0) |
| **Blood disease except for anaemia** | 35 | 1.7% | 119 | 1.5% | 1.2 (0.8-1.7) |
| **Malignancy** | 67 | 3.3% | 149 | 1.8% | 1.8 (1.4-2.4) |
| **Receiving immunosuppressant** | 85 | 4.2% | 224 | 2.3% | 1.5 (1.2-2.0) |
| **Neurological or neuromuscular disease due to immune deficiency** | 24 | 1.2% | 36 | 0.4% | 2.7 (1.6-4.5) |
| **Physical decline associated with a neurological disease or a neuromuscular disease** | 11 | 0.5% | 23 | 0.3% | 1.9 (0.9-3.9) |
| **Chromosomal abnormality** | 3 | 0.2% | 11 | 0.1% | NA |
| **Severe psychosomatic disorder** | 4 | 0.2% | 3 | 0.04% | NA |
| **Sleep apnoea** | 47 | 2.3% | 224 | 2.8% | 0.8 (0.6-1.1) |
| **Obesity** | 232 | 11.4% | 704 | 8.6% | 1.4 (1.2-1.6) |
| **No underlying health condition** | 1095 | 53.7% | 4767 | 58.5% | 0.8 (0.7-0.9) |
| **Smoking status** |  |  |  |  |  |
| Active smoker | 232 | 11.4% | 690 | 8.5% | 1.3 (1.1-1.6) |
| Former smoker | 448 | 22.0% | 1987 | 24.4% | 0.9 (0.8-1.0) |
| Only electronic cigarettes | 41 | 2.0% | 237 | 2.9% | 0.7 (0.5-1.0) |
| Never smoker | 1318 | 64.6% | 5239 | 64.3% |  |
| **Have you had a flu shot within the past year?** | 1039 | 51% | 5998 | 73.6% | 0.4 (0.3-0.4) |
| **Self-reported history of COVID-19** |  |  |  |  |  |
| Yes (I tested positive) | 48 | 0.6% | 10 | 0.5% | 0.8 (0.4-1.7) |
| Yes (I had the symptoms but did not receive a positive test) | 60 | 0.7% | 23 | 1.1% | 1.5 (0.9-2.5) |
| No | 8045 | 80% | 2006 | 98.4% | Ref |
| **Have you had any vaccine side-effects?** |  |  |  |  |  |
| Yes | 331 | 16.2% | 644 | 7.9% | 2.5 (2.2-2.9) |
| No | 1424 | 69.8% | 7051 | 86.5% | Ref |
| Unsure | 284 | 13.9% | 458 | 5.6% | 3.1 (2.6-3.6) |
| **Self-reported overall health** |  |  |  |  |  |
| 1-3 | 40 | 2.0% | 76 | 0.9% | 1.7 (1.2-2.5) |
| 4-6 | 1286 | 63.1% | 4144 | 50.8% | Ref |
| 7-9 | 713 | 35.0% | 3933 | 48.2% | 0.6 (0.5-0.6) |
| **Pregnancy status** |  |  |  |  |  |
| Pregnant | 31 | 1.5% | 36 | 0.4% | 3.3 (2.0-5.3) |
| Not pregnant | 1072 | 52.57 % | 4087 | 50.1% | Ref |
| Desire to be pregnant | 84 | 4.1% | 144 | 1.8% | 2.2 (1.7-2.9) |
| Not applicable | 852 | 41.8% | 3886 | 47.7% | 0.8 (0.8-0.9) |

Ref: reference
NA: Logistic regression was not performed due to too small number (n≤3).

Figure 3 summarizes the reported reasons for vaccine hesitancy or acceptance. The most common reasons for vaccine hesitancy were: “I am worried about vaccine side-effects and/or allergic reactions” (79.7%); followed by, “Many things are not understood about COVID-19 vaccines” (61.8%); and, “I do not think COVID-19 vaccines are safe” (22.2%). The most common reasons for vaccine acceptance were: “I do not want to get infected by COVID-19” (76.0%); followed by, “I do not want to be contagious to others” (74.2%); and, “If I catch COVID-19, I hope that I may only have mild symptoms” (72.5%).

**Figure 3:**
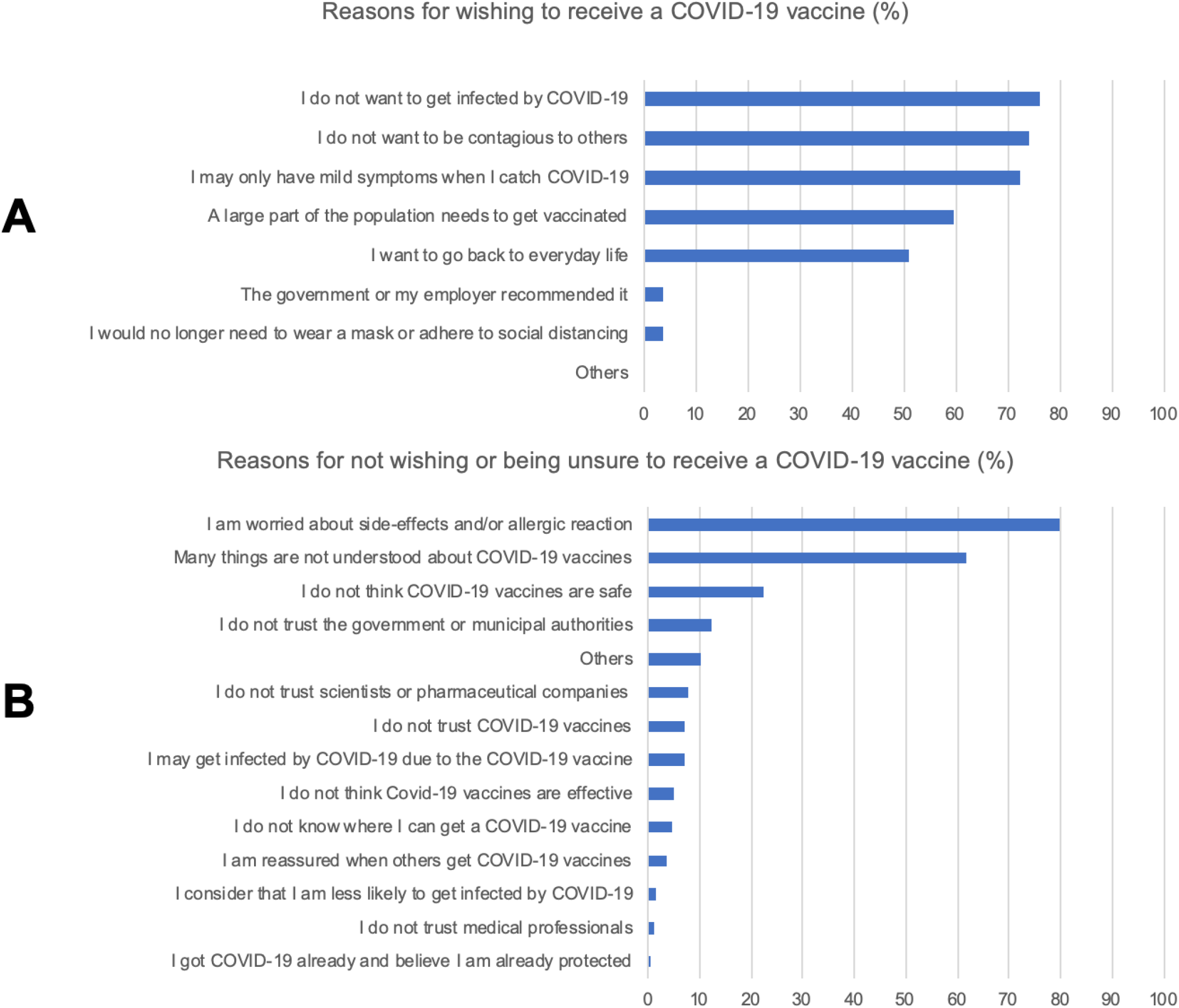
Reasons provided for COVID-19 hesitancy (A) and acceptance (B), COVID-19 vaccine survey, Japan, 2021.

## Discussion

We created a chatbot to answer COVID-19 vaccine FAQs using the most popular messenger app in Japan. Within two months, nearly 60,000 people accessed Corowa-kun. One-fifth of survey participants were COVID-19 vaccine hesitant. The top risk factors for vaccine hesitancy were younger age, female sex, pregnancy or desire for pregnancy, and previous history of a vaccine side-effect. Protective factors included being a physician and a history of having received a flu vaccine the previous season. COVID-19 vaccine acceptance increased after using Corowa-kun. Corowa-kun, a messenger app chatbot, reached the public with COVID-19 vaccine information, helped assess COVID-19 vaccine attitudes, and reassured those with vaccine concerns.

Besides serving as a socialization platform, social media can help reach those in need of health information. Vaccine content is already widely available across social media platforms. [24] However, there is conflicting data regarding the impact of social media on COVID-19 vaccine hesitancy. Wilson et al. revealed that foreign disinformation campaigns are associated with a drop in mean vaccine coverage over time and an increase in negative discussions of vaccines. [24] On the other hand, Ahmed et al. found that people who utilize Twitter and Facebook as sources of health information were more likely to be vaccinated. [25] Furthermore, Ortiz et al. revealed that adolescents who fully engaged with social media health platforms improved their health knowledge, and many were likely to have discussions with others about what they learned. [26] We demonstrated that COVID-19 vaccine acceptance increased after using Corowa-kun. Direct communication between healthcare providers and patients is known to reduce vaccine concerns and improve overall uptake. [27] Public trust in doctors and nurses is relatively high in Japan; 26% reported a lot of trust, and 67% reported some trust. [28] Though there was no direct, real-time communication in Corowa-kun, the content was created by ten physicians, and FAQs were added in response to newly-asked questions by users in the free-text section, which might have contributed to the observed positive change. Healthcare providers may need to become more acquainted with social media and utilize these platforms to reduce vaccine hesitancy with more communication between providers and the public. [29]

Our study found relatively higher rates of COVID-19 vaccine acceptance (80%) compared to previously published studies in Japan. Yoda et al. conducted a cross-sectional study in September 2020 and gathered that 66% of participant were willing to be vaccinated against COVID-19 once a vaccine become available. [30] Another survey performed by Machida et al. in January 2021 noted that 62% of respondents were willing to receive a COVID-19 vaccine. [31] Recently, the U.S. CDC reported that the intent to receive a COVID-19 vaccine increased from 39% in September 2020 to 49% in December 2020. [32] Our survey was conducted in the middle of the COVID-19 fourth wave in Japan, and this may have contributed to the increasing trend of vaccine acceptance. A previous study postulated that women may be more hesitant about COVID-19 vaccines. [33] This could be related to the initial absence of vaccine safety and efficacy data for pregnant women. However, a recent U.S. study showed that among persons who received at least one dose of a COVID-19 vaccine, 63% were female. [34] This discrepancy could be because healthcare workers were offered the vaccine first in the United States, and women account for three-fourths of healthcare workers. Newer evidence suggesting that COVID-19 vaccines are safe in pregnancy may help decrease hesitancy. [20]

Understanding more about vaccine hesitancy could inform public health efforts. Similar to other studies [4] [32], we found lower vaccine acceptance among younger people, those with lower income and education attainment, active smokers, those with a history of a vaccine side-effect, and those with poor self-reported health. We also found that healthcare workers other than physicians were not as protective as physicians were; healthcare workers other than physicians had similar hesitancy levels as non-healthcare workers. Shekhar et al. revealed that only 8% of healthcare workers did not plan to get a vaccine. [35] However, 80% of their responders were providers (e.g., physicians, nurse practitioners), which might have influenced the high acceptance in their study. Healthcare workers other than physicians may have a greater COVID-19 risk than physicians do: nurses and respiratory therapists often have more direct and prolonged patient contact. Special attention to non-physician healthcare workers may be needed, and efforts to understand and address their concerns are critical. Furthermore, people unemployed due to COVID-19 had low vaccine acceptance. Though not common, 12% of vaccine hesitant respondents mentioned distrust of the government or municipal authorities. Japan has a lower level of public trust in the national government compared to other countries: only 4% reported that they had “a lot” of trust in the government. [28] Because of the known association between low vaccine uptake and government distrust [36], authorities may need to adapt their public health responses and messaging to enhance trustworthiness.

Our study has limitations. Only users of the most popular social media/messenger app in Japan (“LINE”) could access Corowa-kun. Our results are not representative of those not using this platform or without access to the internet. Approximately three-fourths of participants were women. Only persons with Japanese literacy could answer the survey. Due to the study design—cross-sectional survey—we could not estimate the incidence of vaccine hesitancy. A hypothetical question (“Would you like to receive a COVID-19 vaccine when possible?”) was used because the COVID-19 vaccine was only available for healthcare professionals and for elderly people in a very limited area at the time that the survey was conducted. Finally, the survey answers may not translate into real-world actions by respondents due to other competing priorities and varying epidemiologic and societal conditions at the time a vaccine is available to them. [37] [38]

## Conclusions

We created Corowa-kun, a free chatbot, to answer COVID-19 vaccine FAQs using the most popular messenger app in Japan. The chatbot was popular, and nearly 10,000 people participated in a cross-sectional survey that helped us identify vaccine hesitancy risk factors. Vaccine hesitancy decreased among Corowa-kun users. Messenger app chatbots may help people obtain appropriate COVID-19 vaccine information in a timely and effective manner.

## Data Availability

The data that support the findings of this study are available from the corresponding author, [T.K], upon reasonable request.

## Conflict of Interest

None

## Acknowledgement

We appreciate Shinji Hirooka, Shinsuke Oyama, Hozumi Kaneko, Makiko Sakamoto, Hiroaki Asahara, Ryosuke Kitano for their technical support for Corowa-kun.

We thank Chaorong Wu and Patrick Ten Eyck for useful discussions around study design. (Institute for Clinical and Translational Science, University of Iowa, Iowa City, Iowa, United States)

## Funding

none

## Notes

### Competing Interest Statement

The authors have declared no competing interest.

### Funding Statement

no funding

### Author Declarations

This study was approved by the institutional review board of Kanto Central Hospital.

