## Supplementary figures and images for "Corowa-kun: Impact of a COVID-19 vaccine information chatbot on vaccine hesitancy, Japan 2021"

### supplemental figure 1

**Supplemental figure 1: Another example for Corowa-kun**

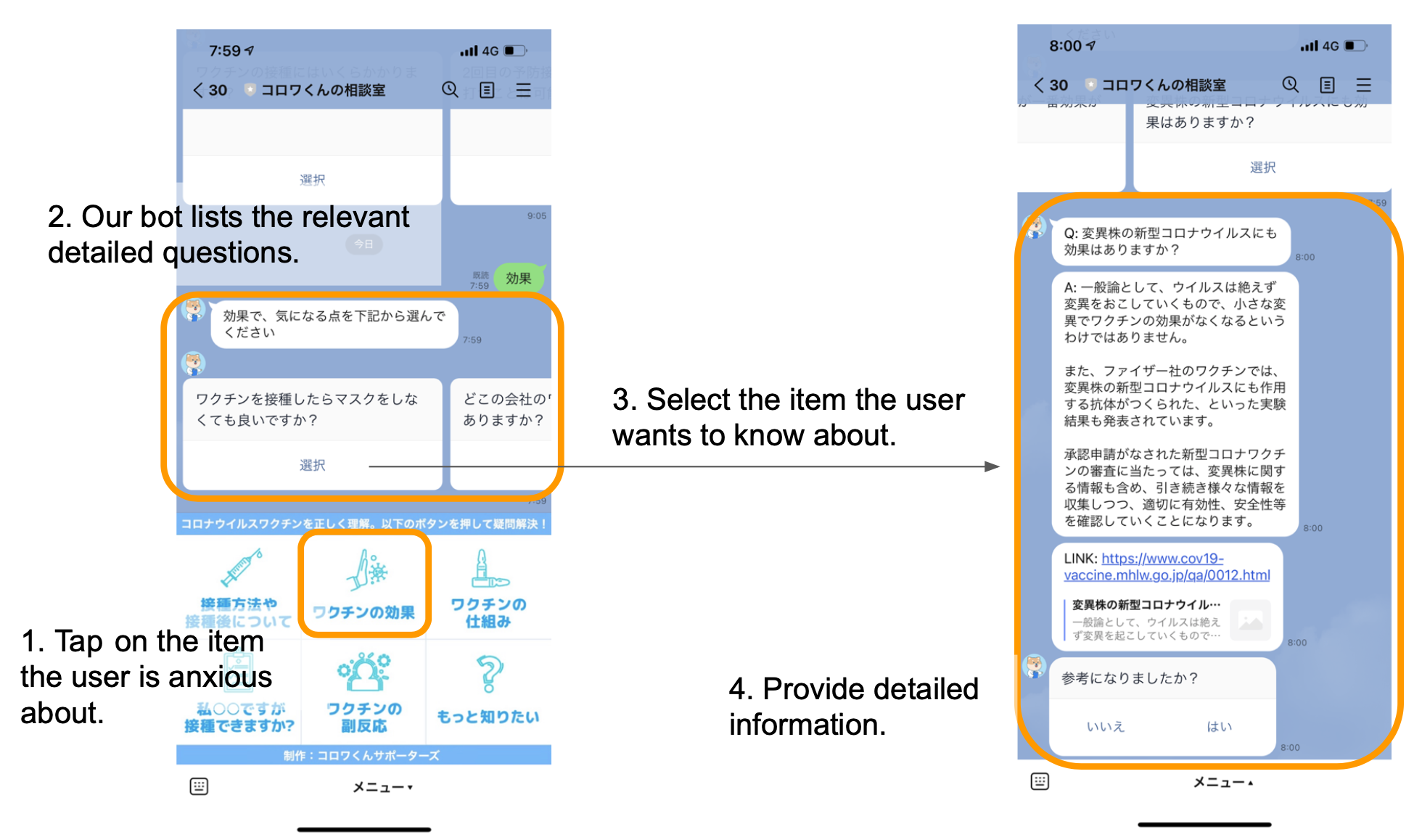

### supplemental figure 2

**Supplemental figure 2: Heat map of Corowa-kun users, Japan, 2021**

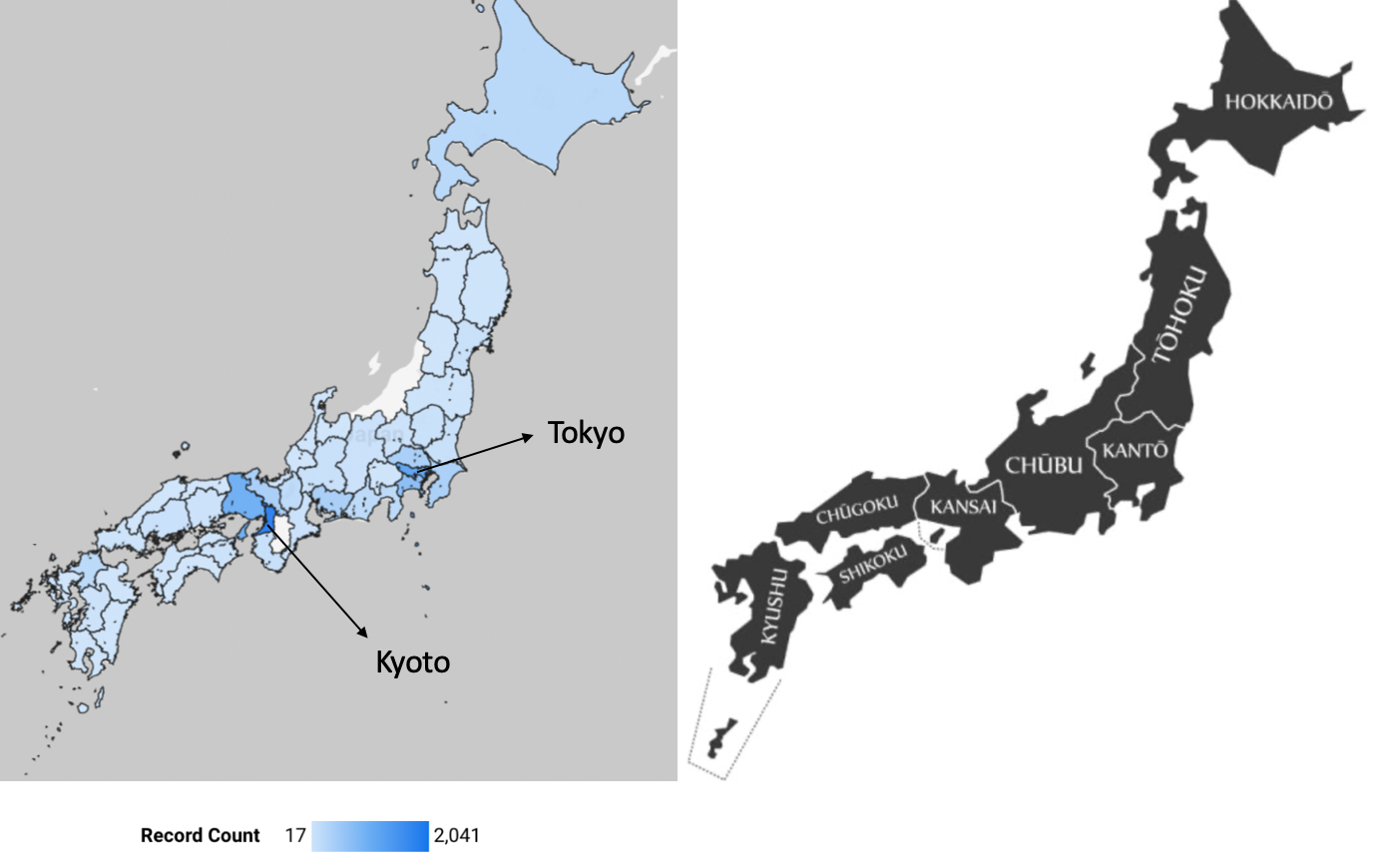
