## supplemental document 1 for "Corowa-kun: Impact of a COVID-19 vaccine information chatbot on vaccine hesitancy, Japan 2021"

**Questionnaire for those who have not yet received a COVID-19 vaccine**

Vaccinations against COVID-19 have begun across the world, including in Japan. “Corowa-kun’s Consultation Room” aims to allay any concerns and resolve any questions you may have regarding COVID-19 vaccines by providing relevant information in an easily digestible and widely available format, delivered via a chatbot on the LINE application. (LINE is a messenger application originally launched in Japan offering similar services to other applications like WhatsApp.)

Using this questionnaire, we hope to gain insights into Japanese people’s thoughts about and understanding of COVID-19 vaccines, so that we can, in turn, improve the service offered by “Corowa-kun’s Consultation Room”.

There are 22 questions which will take you approximately 5 minutes to complete. Any information provided on this form is confidential, and we comply with applicable legislation to ensure your data is protected from unauthorised access or leakage. The information on this form shall only be used with a view to improving the service offered by “Corowa-kun’s Consultation Room”. None of your data shall be provided to any third-party without your express prior consent. For your information, our privacy policy is here: <https://corowakun-supporters.studio.site/privacypolicy/(Japanese)>.

**Screening questions**

1. Are you at least 16 years old?

- Yes
- No

2. Have you received a COVID-19 vaccine yet?

- Yes, I have
- No, I have not yet

3. Do you consent to the above and to proceed with the questionnaire?

- Agree
- Disagree

**Questionnaire**

1. How old are you?

(Select a number from 16 to 100)

2. What is your sex?
a. Male
b. Female
c. Other
d. No answer
3. In which prefecture do you live?

(Select from 47 prefectures in Japan + “abroad”)

4. What’s the highest level of education you have attained?
a. Elementary school and junior high school
b. High school
c. College or professional school
d. Undergraduate school (Bachelor’s degree)
e. Postgraduate school (Master’s or Doctoral degree)

5. Are you currently employed (whether under a permanent contract or on a consulting basis)?
a. Full-time
b. Part-time
c. Student
d. Retired
e. Homemaker
f. Unemployed (due to COVID-19)
g. Unemployed (due to non-COVID-19 reasons)
6. What is your current occupation? Please select one of the below.
a. Medical doctor in a health care facility
b. Healthcare professional in a health care facility (other than a medical doctor)
c. Other form of work in a health care facility (e.g. as a receptionist, cleaner, etc.)
d. Healthcare professional in a nursing or welfare facility
e. Engaged in an educational facility (e.g. nursery, elementary school, high school or university)
f. Work in a restaurant or supermarket
g. Work in transportation or delivery (e.g. bus, train, or taxi driver, or postal carrier)
h. Paramedic
i. Police officer
j. Work in an office (excluding any of the other options in this question)
k. Self-employed
l. Other
7. Are you married?
a. Married
b. Never married
c. Divorced
8. Are you pregnant?
a. Pregnant
b. Not pregnant
c. Desire to be pregnant
9. Do you live with a child under the age of 16?
a. Yes
b. No
10. Do you live with a person aged 65 or over?
a. Yes
b. No
11. How many people do you live with, including you?

(Select from 1-6, and more than 6)

12. What is your income?
a. Less than JPY 2 million
b. JPY 200 million < JPY 400 million
c. JPY 400 million < JPY 600 million
d. JPY 600 million < JPY 800 million
e. JPY 800 million or higher
13. Do you have any of the following illnesses / diseases? (Select all that apply)
a. Chronic respiratory disease
b. Chronic heart disease (including hypertension)
c. Chronic kidney disease
d. Chronic liver disease (except for fatty liver or chronic hepatitis)
e. Diabetes treated with insulin or other oral medication, or diabetes with any complication
f. Blood disease (except for anaemia)
g. Disease with an immune suppression (including neoplasms regardless of treatment)
h. Receiving treatment that may suppress the immune system e.g. steroids
i. Neurological or neuromuscular disease due to immune deficiency
j. Physical decline associated with a neurological disease or a neuromuscular disease (such as a respiratory disease)
k. Chromosomal abnormality
l. Sever psychosomatic disorder (overlapping severe physical disability and severe intellectual disability)
m. Sleep apnea
n. Obesity (BMI over 30, which would typically be a person of 170cm height weighing 87kg, or 160cm height weighing 77kg)
o. No underlying health conditions
p. Other
14. Do you smoke?
a. Yes, I’m a smoker
b. I quit smoking
c. I smoke only electronic cigarettes
d. I have never smoked
15. Did you receive a flu vaccine between autumn 2020 and spring 2021?
a. Yes
b. No
16. Have you ever experienced a side-effect or allergy after a vaccination?
a. Yes
b. No
c. I do not know
17. How do you rate your current health status? (Answer between 1 to 9, where 1= very poor, 5 = normal and 9 = very good)
18. Have you ever caught COVID-19?
a. Yes (I tested positive)
b. Yes (I had the symptoms but did not receive a positive test)
c. No
19. Would you like to receive a COVID-19 vaccine when possible?
a. Yes
b. No
c. I do not know

*If a respondent answers “Yes” to Q19*

20. What is the reason that you would like to get a COVID-19 vaccine?
a. I do not want to get infected by COVID-19
b. If I catch COVID-19 I hope that I may only have mild symptoms
c. I do not want to be contagious to others
d. I want to go back to everyday life before the COVID-19 pandemic
e. I hope that I would no longer need to wear a mask or adhere to social distancing after vaccination
f. The government or my employer recommended it
g. A large part of the population need to get vaccinated
h. Other
21. Did your view on COVID-19 vaccines change after you used “Corowa-kun’s Consultation Room”?

|  | a. I want to receive a COVID-19 vaccine | b. I don’t want to receive a COVID-19 vaccine | c. No opinion either way |
| --- | --- | --- | --- |
| Before |  |  |  |
| After | The same answer of question #19 used | | |

*If a respondent answers “No” to Q19*

20. What is the reason that you do not wish to receive a COVID-19 vaccine?

a. I got COVID-19 already and believe I am already protected
b. I may get infected by COVID-19 due to the COVID-19 vaccine
c. I am worried about any side effect and/ or allergic reaction to the vaccine
d. I do not think COVID-19 vaccines are safe
e. I do not think COVID-19 vaccines are effective
f. Many things are not understood about COVID-19 vaccines
g. I do not trust COVID-19 vaccines
h. I consider that I am less likely to get infected by COVID-19
i. I am reassured when others get COVID-19 vaccines
j. I do not know where I can get a COVID-19 vaccine
k. I do not trust the government or municipal authorities
l. I do not trust scientists or pharmaceutical companies involved in COVID-19 vaccines
m. I do not trust medical professionals
n. Other
21. Did your view on COVID-19 vaccines change after you used “Corowa-kun’s Consultation Room”?

|  | a. I want to receive a COVID-19 vaccine | b. I don’t want to receive a COVID-19 vaccine | c. No opinion either way |
| --- | --- | --- | --- |
| Before |  |  |  |
| After | The same answer of question #19 used | | |
